# Latent tuberculosis and depressive symptoms in household contacts of persons with active TB: A cohort study

**DOI:** 10.1101/2022.11.15.22282271

**Authors:** Jerome Galea, Alexander L. Chu, Annika Sweetland, Judith Jimenez, Rosa Yataco, Roger Calderón, Zibiao Zhang, Chuan-Chin Huang, Leonid Lecca, Megan Murray

**Author notes:** **Corresponding Author Contact:** Jerome T Galea, 4202 E Fowler Avenue, MHC 1400, Tampa, FL, USA 33620.

## Abstract

**Background:** Depression is common among persons with TB and associated with poor clinical outcomes. However, little is known about the relationship between latent TB infection (LTBI) and depression. Here, we assessed the association between LTBI and depressive symptoms among household contacts (HHCs) of patients receiving TB treatment.

**Methods:** We enrolled 1,009 HHCs of 307 patients receiving TB treatment in Lima, Peru, 2016-2018. We assessed HHC LTBI status at enrollment using interferon gamma release assay (IGRA). Depressive symptoms were assessed at baseline and 12 months later using the Patient Health Questionnaire-9 (PHQ-9) using a cutoff of ≥5. We used logistic regression to estimate the odds ratio for PHQ-9 ≥5 comparing HHCs with and without baseline LTBI.

**Results:** Among 921 HHCs, 378 (41.0%) had LTBI at baseline, and 70 (12.4%) of 563 HHCs had PHQ-9 ≥5. Compared to HHCs without LTBI at enrollment, those with LTBI had almost two times the odds of PHQ-9 ≥5 at follow-up after controlling for potential confounders (adjusted OR, 1.93, 95% CI, 1.09-3.39); this association was driven by greater severities of depressive symptoms.

**Conclusion:** HHCs with LTBI had increased odds of depressive symptoms one year later. This at-risk population may benefit from mental health screening and interventions integrated within TB programs.

## INTRODUCTION

Tuberculosis (TB) and depression are among the leading causes of death and disability worldwide and commonly co-occur.^1–4^ Individuals with active TB have more than three times the risk of developing depression than members of the general population, and depression may affect as many as half of all patients receiving TB treatment.^5^ Importantly, depression is associated with 2.85 times the risk for TB-related death and 8.70 times the risk for loss to follow up from TB treatment.^6^ Consequently, most mental health research has focused on the urgent need of integrating mental health services for people with active TB,^7^ and the World Health Organization (WHO) now includes screening and management of mental health comorbidities in its 2022 guidance for national strategic planning for TB.^8^

While emphasis on the mental health of people with active TB disease is appropriate, the association between common mental disorders like depression and latent TB infection (LTBI)—TB infection that presents with no clinical symptoms—also warrants investigation. It has been estimated that about 10% of individuals with LTBI progress to active TB,^9^ and a wide range of factors associated with progression include Human Immunodeficiency Virus (HIV) infection, cigarette smoking, and medical comorbidities like diabetes mellitus^10^; however, they do not typically include common mental disorders that have been associated with compromised immune function like depression. A recent systematic review evaluating the health-related quality of life in individuals with LTBI (comprising of nine studies and 8,086 individuals) found that people with LTBI reported poorer measures of mental health compared with those without LTBI. However, only two studies specifically measured depression, and none were based in high TB burden countries.^11^

Few studies have considered mental disorders among close contacts of persons with TB, an at-risk group for LTBI and active TB. Preliminary evidence from Taiwan suggests that female contacts have an increased risk of developing depression within the first two years after TB diagnosis of someone in their household compared with matched female controls in the general population.^12^ However, it remains unclear if the mental health risks among contacts of persons with TB are due to biological factors related to LTBI and/or psychosocial factors like stigma and caregiving-related stressors.^12–15^

Here, we report the association between LTBI and depressive symptoms in household contacts (HHCs) of persons with TB in Lima, Peru.

## METHODS

### Study Setting and Design

This was a secondary analysis of data collected from a larger prospective cohort study that investigated the metabolic factors that control the spectrum of human TB.^16^ Briefly, 350 persons aged ≥14 years with newly diagnosed active TB and living with at least two HHCs were consecutively recruited from participating health centers in Metropolitan Lima and subsequently followed during 2016-2021. Next, we invited 1,050 of their HHCs to participate in a study with a scheduled one-year follow-up visit and unscheduled follow-up visits beforehand for those who decided to voluntarily withdraw from the study or developed active TB. At the time of HHC enrollment, we collected information on HHC socio-demographics (age and sex), health-related behaviors (smoking and drinking statuses), nutritional status, current use of isoniazid preventive therapy (IPT), HIV status, medical comorbidities (heart disease, diabetes mellitus, hypertension, asthma, and renal disease), and socioeconomic status (SES).

### Classification of Household Contact Latent TB Infection Status

HHC LTBI status was assessed at time of enrollment using the Interferon-Gamma Release Assays (IGRA, QuantiFERON-TB Gold or QuantiFERON-TB Gold Plus, QIAGEN Germantown, MD). These tests were conducted per manufacturer instructions. Briefly, no less than 4 mL of blood were collected from each participant and then transported to the laboratory within the same day. A total of 1 mL of the blood was placed into four blood collection tubes: a negative control, a positive control (T cell mitogen), TB1 (ESAT-6 and CFP-10 peptides), and TB2 (ESAT-6 and CFP-10 peptides). Of note, the QuantiFERON-TB Gold only utilized the TB1 tube. Following a 16-to 24-hour incubation period at 37°C, the plasma was separated after the tubes underwent centrifugation at 2500×g for 15 minutes. The concentration of interferon gamma was determined by enzyme-linked immunosorbent assay, and the results were interpreted based on the manufacturer’s pre-determined cut-off value. A positive IGRA result was determined when the values of TB1 or TB2 minus Nil were ≥0.35 IU/ml. HHCs without clinical symptoms of TB and had positive IGRA results were considered to have LTBI.

### Study Outcomes

We used the Patient Health Questionnaire-9 (PHQ-9) to assess depressive symptoms among HHCs (17). This tool assesses both the number and frequency of nine depressive symptoms (loss of interest or pleasure, depressed mood, sleeping difficulties, fatigue, poor appetite, self-guilt, concentration difficulties, psychomotor changes, and suicidal ideation) over the preceding two weeks on a scale from 0 (i.e., experiencing no depressive symptoms) to 27 (i.e., experiencing all depressive symptoms nearly every day).^17,18^ We considered individuals to have depressive symptoms at follow-up if they had PHQ-9 scores 5-27 with PHQ-9 scores 5-9 indicating mild depression, PHQ-9 scores 10-14 for moderate, PHQ-9 scores 15-19 for moderately severe, and PHQ-9 scores 20-27 for severe. We considered all HHCs with PHQ-9 data collected at 12 months with a two-month data collection buffer period (i.e., 10-14 months). Because the PHQ-9 only assesses depressive symptoms over a two-week period, we sought to expand the period by which baseline LTBI may affect depressive symptoms one year later.

### Statistical Analysis

We conducted our analysis on HHCs ≥12 years of age, because the PHQ-9 was not originally designed to assess depressive symptoms among children. We used logistic regression to estimate the odds ratio (OR) and 95% confidence intervals (CIs) for depressive symptoms, defined by PHQ-9 ≥5. We first performed bivariate analyses for covariates that may confound or modify any association between baseline LTBI and depressive symptoms at follow-up. We chose the following covariates based on a priori background knowledge: sex, age, smoking status, drinking status, nutritional status, current use IPT, HIV status, status of medical co-morbidities (heart disease, hypertension, asthma, renal disease, and diabetes), and SES. These covariates were entered into a backwards stepwise algorithm to construct the multivariate models, and we retained the covariates with p<0.1 in our final multivariate models. For any covariate with p<0.1, we removed them from the multivariate model if its prevalence in the cohort was <3%. We used complete data for all our multivariate analyses.

To explore the possibility that the effect of LTBI at baseline may vary for different severities of depressive symptoms at follow-up, we first repeated the main analysis but only compared HHCs with PHQ-9 scores 5-9 (mild symptoms) with those with PHQ-9 scores 0-4 (no/minimal symptoms). We then repeated the same analysis, comparing only HHCs with PHQ-9 scores 10-27 (moderate, moderately severe, or severe symptoms) and those with PHQ-9 scores 0-4.

We performed three sensitivity analyses. First, to determine if depressive symptoms were limited to HHCs who developed TB disease during follow-up, we further adjusted our multivariate model for HHC incident TB disease status. Second, to assess if our findings were due to recently or distantly acquired LTBI, we restricted our analysis to HHCs with IGRA results available for both baseline and 12 months of follow-up. We considered HHCs to have acquired recent LTBI if they had converted their IGRA status from negative to positive during follow-up. We then assessed the effect of recently acquired LTBI on depressive symptoms at follow-up. Third, to assess whether our findings were sensitive to the choice of PHQ-9 score cutoff in defining depressive symptoms, we repeated the main analysis using PHQ-9 ≥10 to define depressive symptoms. All statistical analyses were performed using R Version 3.6.2 (R Foundation for Statistical Computing, Vienna, Austria).

### Ethics Statement

The research ethics committee of the National Institute of Health of Peru (protocol OEE-004-16) and Harvard Medical School (protocol #IRB16-1173) reviewed and approved the original study prior to implementation.

## RESULTS

### Household Contact Characteristics and Bivariate Analyses

We recruited 1,009 HHCs of 307 index patients with culture-positive pulmonary TB of whom 921 were ≥12 years old (Figure 1). At baseline, 378 (41.2%) of 918 HHCs were considered to have LTBI. During follow-up, 70 (12.4%) of 563 HHCs had PHQ-9 ≥5. Baseline LTBI status was associated with older HHC age and being overweight (Table 1).

**Table 1.**
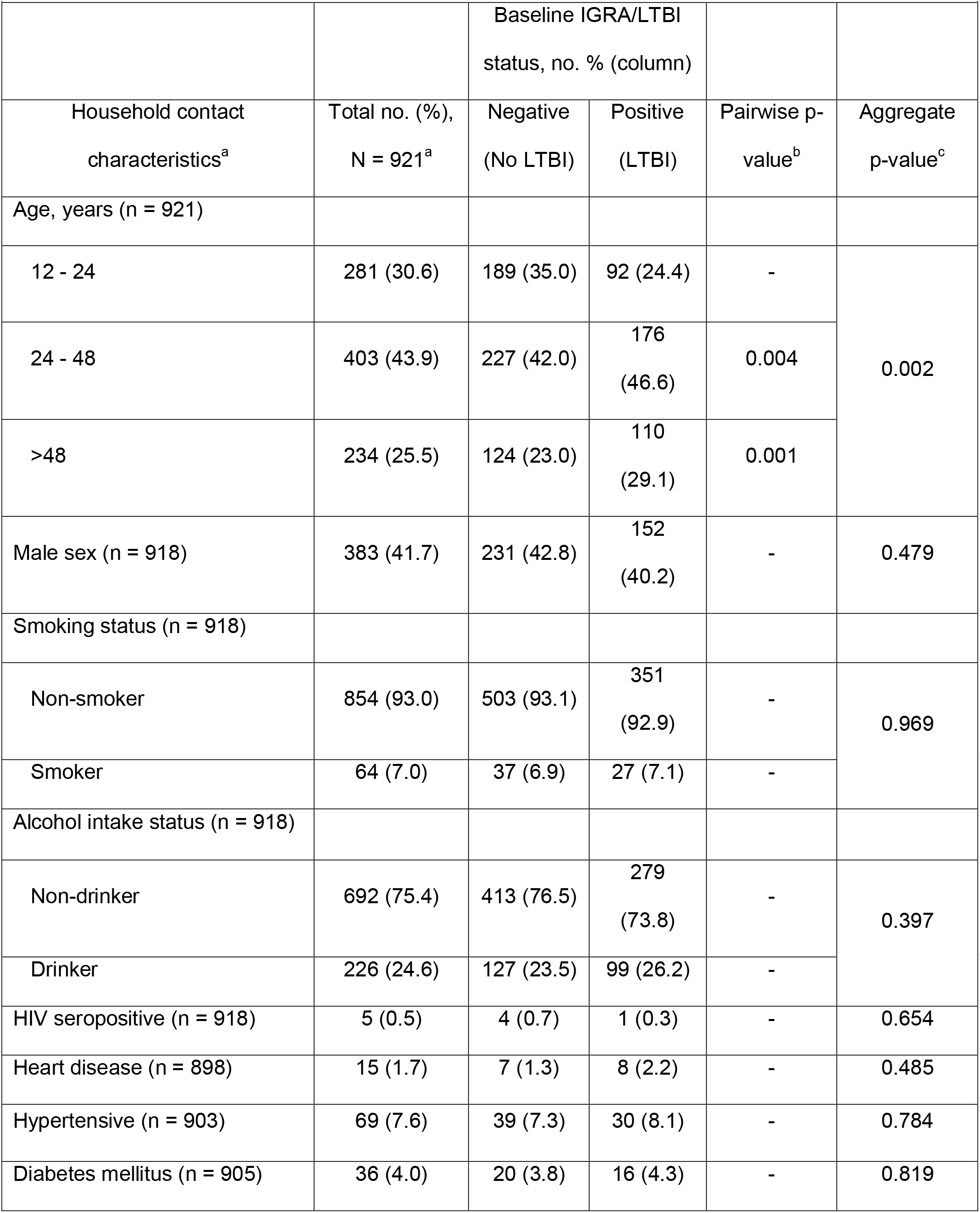
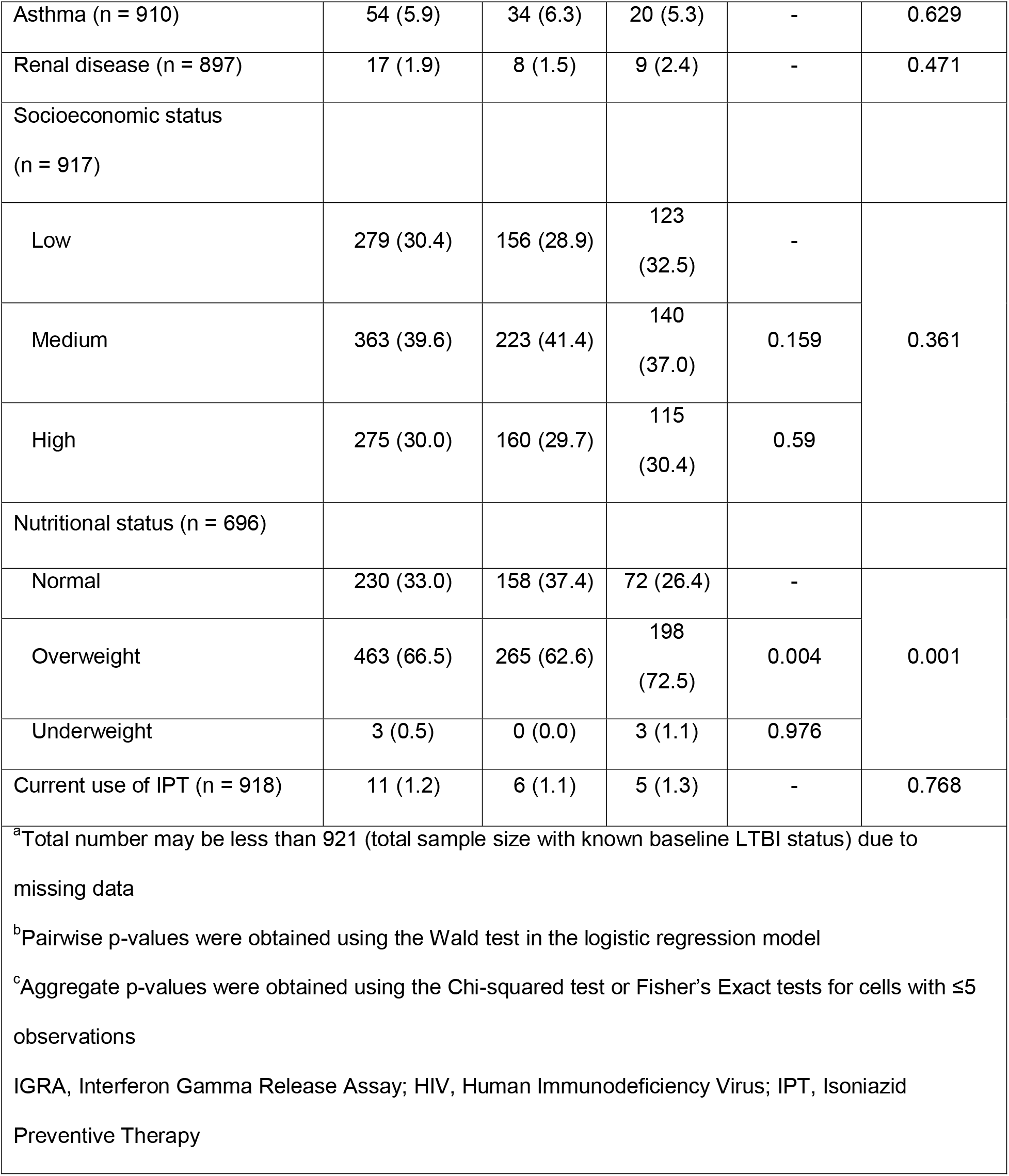
Household contact baseline characteristics, by baseline IGRA status (N = 921)

|  |  |  |  |  |  |
| --- | --- | --- | --- | --- | --- |
|  |  | Baseline IGRA/LTBI status, no. % (column) |  |  |  |
| Household contact characteristics<sup>a</sup> | Total no. (%), N = 921<sup>a</sup> | Negative (No LTBI) | Positive (LTBI) | Pairwise p-value<sup>b</sup> | Aggregate p-value<sup>c</sup> |
| Age, years (n = 921) |  |  |  |  |  |
| 12 - 24 | 281 (30.6) | 189 (35.0) | 92 (24.4) | - | 0.002 |
| 24 - 48 | 403 (43.9) | 227 (42.0) | 176 (46.6) | 0.004 |  |
| >48 | 234 (25.5) | 124 (23.0) | 110 (29.1) | 0.001 |  |
| Male sex (n = 918) | 383 (41.7) | 231 (42.8) | 152 (40.2) | - | 0.479 |
| Smoking status (n = 918) |  |  |  |  |  |
| Non-smoker | 854 (93.0) | 503 (93.1) | 351 (92.9) | - | 0.969 |
| Smoker | 64 (7.0) | 37 (6.9) | 27 (7.1) | - |  |
| Alcohol intake status (n = 918) |  |  |  |  |  |
| Non-drinker | 692 (75.4) | 413 (76.5) | 279 (73.8) | - | 0.397 |
| Drinker | 226 (24.6) | 127 (23.5) | 99 (26.2) | - |  |
| HIV seropositive (n = 918) | 5 (0.5) | 4 (0.7) | 1 (0.3) | - | 0.654 |
| Heart disease (n = 898) | 15 (1.7) | 7 (1.3) | 8 (2.2) | - | 0.485 |
| Hypertensive (n = 903) | 69 (7.6) | 39 (7.3) | 30 (8.1) | - | 0.784 |
| Diabetes mellitus (n = 905) | 36 (4.0) | 20 (3.8) | 16 (4.3) | - | 0.819 |
| Asthma (n = 910) | 54 (5.9) | 34 (6.3) | 20 (5.3) | - | 0.629 |
| Renal disease (n = 897) | 17 (1.9) | 8 (1.5) | 9 (2.4) | - | 0.471 |
| Socioeconomic status<br>(n = 917) |  |  |  |  |  |
| Low | 279 (30.4) | 156 (28.9) | 123<br>(32.5) | - | 0.361 |
| Medium | 363 (39.6) | 223 (41.4) | 140<br>(37.0) | 0.159 |  |
| High | 275 (30.0) | 160 (29.7) | 115<br>(30.4) | 0.59 |  |
| Nutritional status (n = 696) |  |  |  |  |  |
| Normal | 230 (33.0) | 158 (37.4) | 72 (26.4) | - | 0.001 |
| Overweight | 463 (66.5) | 265 (62.6) | 198<br>(72.5) | 0.004 |  |
| Underweight | 3 (0.5) | 0 (0.0) | 3 (1.1) | 0.976 |  |
| Current use of IPT (n = 918) | 11 (1.2) | 6 (1.1) | 5 (1.3) | - | 0.768 |
| <sup>a</sup>Total number may be less than 921 (total sample size with known baseline LTBI status) due to missing data<br><sup>b</sup>Pairwise p-values were obtained using the Wald test in the logistic regression model<br><sup>c</sup>Aggregate p-values were obtained using the Chi-squared test or Fisher's Exact tests for cells with $\leq 5$ observations<br>IGRA, Interferon Gamma Release Assay; HIV, Human Immunodeficiency Virus; IPT, Isoniazid Preventive Therapy | | | | | |

**Figure 1.**
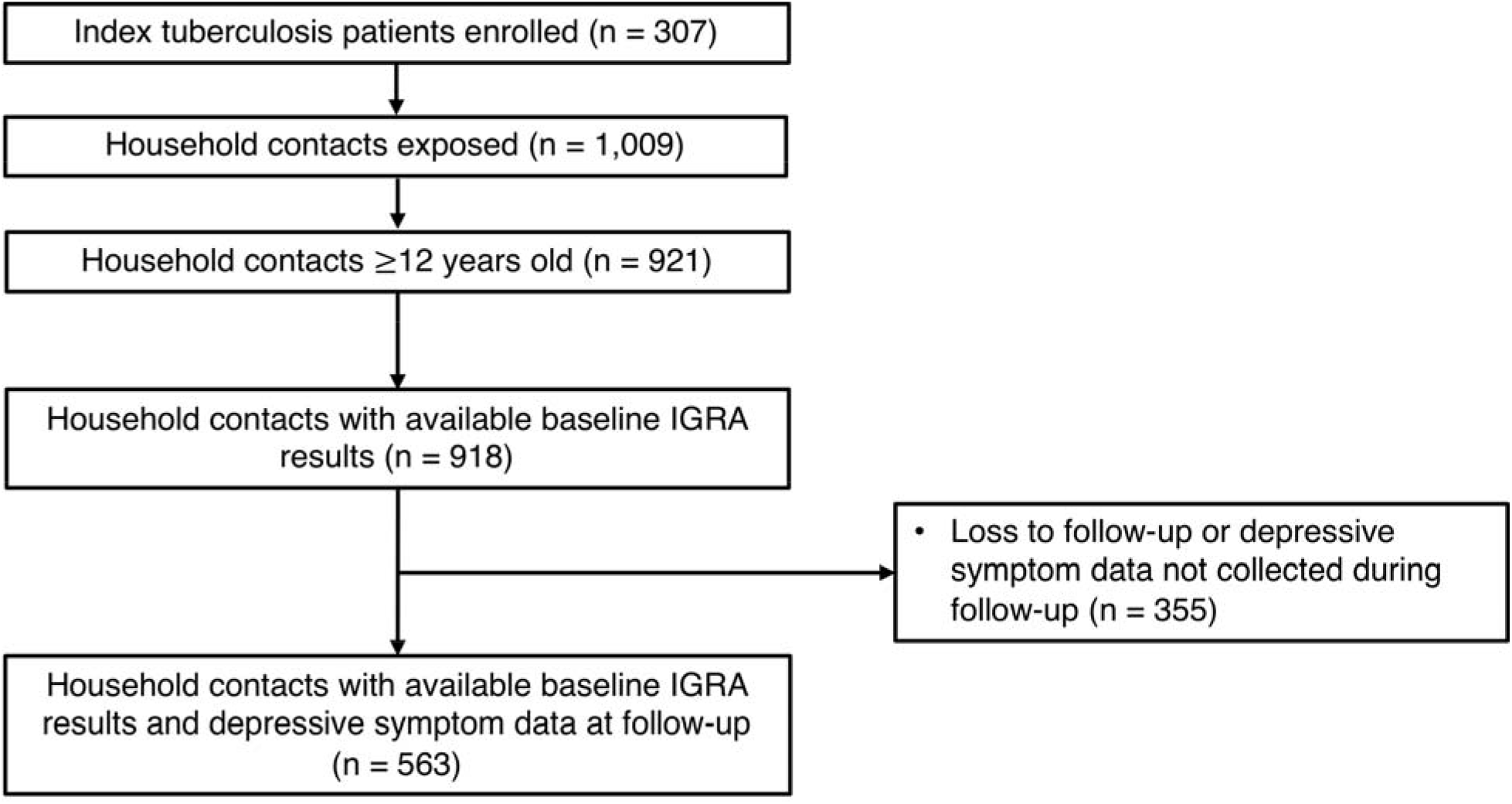
Study flow chart for index patients receiving TB treatment and household contacts.

### Comparison of Baseline Characteristics of Missing and Analytic Samples

By time of follow-up, 358 (38.9%) of 921 HHCs had missing PHQ-9 data. However, we did not find any statistically significant differences in HHC characteristics between the complete/analytic (i.e., HHCs with baseline IGRA results and PHQ-9 data at follow-up) and missing (i.e., HHCs with only baseline IGRA results) samples (Table S1).

### Baseline Latent TB Infection and Depressive Symptoms at Follow-up

A total of 563 (61.1%) of 921 HHCs had available PHQ-9 data collected during follow-up, and 70 (12.4%) were found to have PHQ-9 ≥5. Depressive symptoms at follow-up were associated with HHC female sex and renal disease (Table 2). After adjusting for potential confounders, HHCs with LTBI at baseline had more than twice the odds of experiencing depressive symptoms at follow-up compared with those without LTBI (adjusted OR, 2.07, 95% CI, 1.06–3.95) (Table 3). Our results did not change substantially when we further adjusted for incident TB disease status (adjusted OR, 1.93, 95% CI, 1.09–3.39) (Table 3). We observed similar trends when we restricted our analysis to HHCs who recently acquired LTBI during follow-up; however, the sample size was too small to make any definitive conclusions (Table S2).

**Table 2.** Household contact baseline characteristics, by depressive symptom status at 12 months of follow-up (N = 563)

|  |  |  |  |  |  |
| --- | --- | --- | --- | --- | --- |
|  |  | Depressive symptom status<br>at 12 months of follow-up, no.<br>% (row) |  |  |  |
| Household contact<br>characteristics<sup>a</sup> | Total no. (%),<br>N = 563<sup>a</sup> | PHQ-9<br>scores 0-4 | PHQ-9<br>scores 5-27 | Pairwise p-value<sup>b</sup> | Aggregate p-value<sup>c</sup> |
| Age, years (n = 563) |  |  |  |  |  |
| 12 - 24 | 182 (32.3) | 159 (87.4) | 23 (12.6) |  | 0.929 |
| 24 - 48 | 242 (43.0) | 211 (88.5) | 31 (11.5) | 0.958 |  |
| >48 | 139 (24.7) | 123 (87.2) | 16 (12.8) | 0.76 |  |
| Male sex (n = 563) | 237 (42.1) | 222 (93.7) | 15 (6.3) |  | 0.000 |
| Smoking status<br>(n = 563) |  |  |  |  |  |
| Non-smoker | 522 (92.7) | 457 (87.5) | 65 (12.5) |  | 1.000 |
| Smoker | 41 (7.3) | 36 (87.8) | 5 (12.2) |  |  |
| Alcohol intake status<br>(n = 563) |  |  |  |  |  |
| Non-drinker | 422 (75.0) | 128 (90.8) | 13 (9.2) |  | 0.235 |
| Drinker | 141 (25.0) | 365 (86.5) | 57 (13.5) |  |  |
| HIV seropositive<br>(n = 563) | 3 (0.5) | 3 (100.0) | 0 (0.0) |  | 1.000 |
| Heart disease<br>(n = 546) | 11 (2.0) | 9 (81.8) | 2 (18.2) |  | 0.376 |
| Hypertensive (n = 550) | 47 (8.5) | 41 (87.2) | 6 (12.8) |  | 1.000 |
| Diabetes mellitus<br>(n = 553) | 22 (4.0) | 20 (90.9) | 2 (9.1) |  | 1.000 |
| Asthma (n = 560) | 35 (6.2) | 29 (82.9) | 6 (17.1) |  | 0.425 |
| Renal disease<br>(n = 550) | 12 (2.2) | 8 (66.7) | 4 (33.3) |  | 0.045 |
| Socioeconomic status<br>(n = 563) |  |  |  |  |  |
| Low | 173 (30.7) | 147 (85.0) | 26 (15.0) |  | 0.108 |
| Medium | 217 (38.5) | 187 (86.2) | 30 (13.8) | 0.736 |  |
| High | 173 (30.7) | 159 (91.9) | 14 (8.1) | 0.047 |  |
| Nutritional status<br>(n = 516) |  |  |  |  |  |
| Normal | 182 (35.3) | 163 (89.6) | 19 (10.4) |  | 0.409 |
| Overweight | 332 (64.3) | 284 (85.5) | 48 (14.5) | 0.198 |  |
| Underweight | 2 (0.4) | 2 (100.0) | 0 (0.0) | 0.984 |  |
| Current use of IPT<br>(n = 563) | 8 (1.4) | 6 (75.0) | 2 (25.0) |  | 0.261 |
| <sup>a</sup>Total number may be less than 563 (total sample size for HHCs with known baseline LTBI status and depressive symptom status at 12 months of follow-up) due to missing data<br><sup>b</sup>Pairwise p-values were obtained using the Wald test in the logistic regression model<br><sup>c</sup>Aggregate p-values were obtained using the Chi-squared test or Fisher's Exact tests for cells with $\leq 5$ observations<br>HIV, Human Immunodeficiency Virus; IPT, Isoniazid Preventive Therapy | | | | | |

**Table 3.** Association between baseline LTBI and depressive symptoms at 12 months of follow-up among HHCs of patients receiving TB treatment.

|  | Depressive symptom status at 12 months of follow-up<sup>a</sup>, no. % (row) |  | Univariate (N = 563) | Multivariate (N = 479)<sup>b</sup> | Multivariate - further adjusted for incident tuberculosis disease status (N = 516)<sup>c</sup> |
| --- | --- | --- | --- | --- | --- |
| Baseline IGRA/LTBI status | PHQ-9 scores 0-4 | PHQ-9 scores 5-27 | Crude OR (95% CI) | Adjusted OR (95% CI) | Adjusted OR (95% CI) |
| Negative (No LTBI) | 378 (89.6) | 44 (10.4) | Reference | Reference | Reference |
| Positive (LTBI) | 115 (81.6) | 26 (18.4) | 1.94 (1.13 - 3.27) | 2.07 (1.06 - 3.95) | 1.93 (1.08 - 3.39) |

<sup>a</sup>Depressive symptoms were defined as PHQ-9 = 5-9 among household contacts with available PHQ-9 score at 12 months of follow-up
<sup>b</sup>Adjusted for household contact sex, age, smoking status, drinking status, nutritional status, isoniazid preventive therapy status, HIV status, heart disease status, hypertension status, asthmatic status, renal disease status, diabetic status, and socioeconomic status
<sup>c</sup>Adjusted for household contact sex, age, socioeconomic status, nutritional status, isoniazid preventive therapy, and incident tuberculosis disease status (final model)
LTBI, Latent Tuberculosis Infection; IGRA, Interferon Gamma Release Assay; OR, Odds Ratio; CI, Confidence Interval

When we used PHQ-9 ≥10 to define depressive symptoms, we found that baseline LTBI was more strongly associated with depressive symptoms at follow-up (adjusted OR, 4.30, 95% CI, 1.61-11.90) (Table S3). This finding was confirmed when we conducted two separate analyses comparing PHQ-9 scores 10-27 vs. 0-4 with PHQ-9 scores 5-9 vs. 0-4 (adjusted OR, 4.49, 95% CI, 1.68-12.37 for PHQ-9 scores 10-27 vs. 0-4; adjusted OR, 1.35, 95% CI, 0.66-2.63 for PHQ-9 scores 5-9 vs. 0-4; Tables 4A/B).

**Table 4A.** Association between baseline LTBI and depressive symptoms at 12 months of follow-up among HHCs with PHQ-9 scores 5-9 vs. 0-4.

|  | Depressive symptom status at 12 months of follow-up<sup>a</sup>, no. %<br>(row) |  | Univariate<br>(N = 544) | Multivariate<br>(N = 498)<sup>b</sup> |
| --- | --- | --- | --- | --- |
| Baseline IGRA/LTBI status | PHQ-9 scores 0-4 | PHQ-9 scores 5-9 | Crude OR (95% CI) | Adjusted OR (95% CI) |
| Negative (No LTBI) | 378 (91.5) | 35 (8.5) | Reference | Reference |
| Positive (LTBI) | 115 (87.8) | 16 (12.2) | 1.50 (0.78 - 2.77) | 1.35 (0.66 - 2.63) |

<sup>a</sup>Depressive symptoms were defined as PHQ-9 scores 5-9 among household contacts with available PHQ-9 scores at 12 months of follow-up
<sup>b</sup>Adjusted for HHC sex, age (categorical), SES, nutritional status, and current use of isoniazid preventive therapy
LTBI, Latent Tuberculosis Infection; IGRA, Interferon Gamma Release Assay; OR, Odds Ratio; CI, Confidence Interval

**Table 4B.**
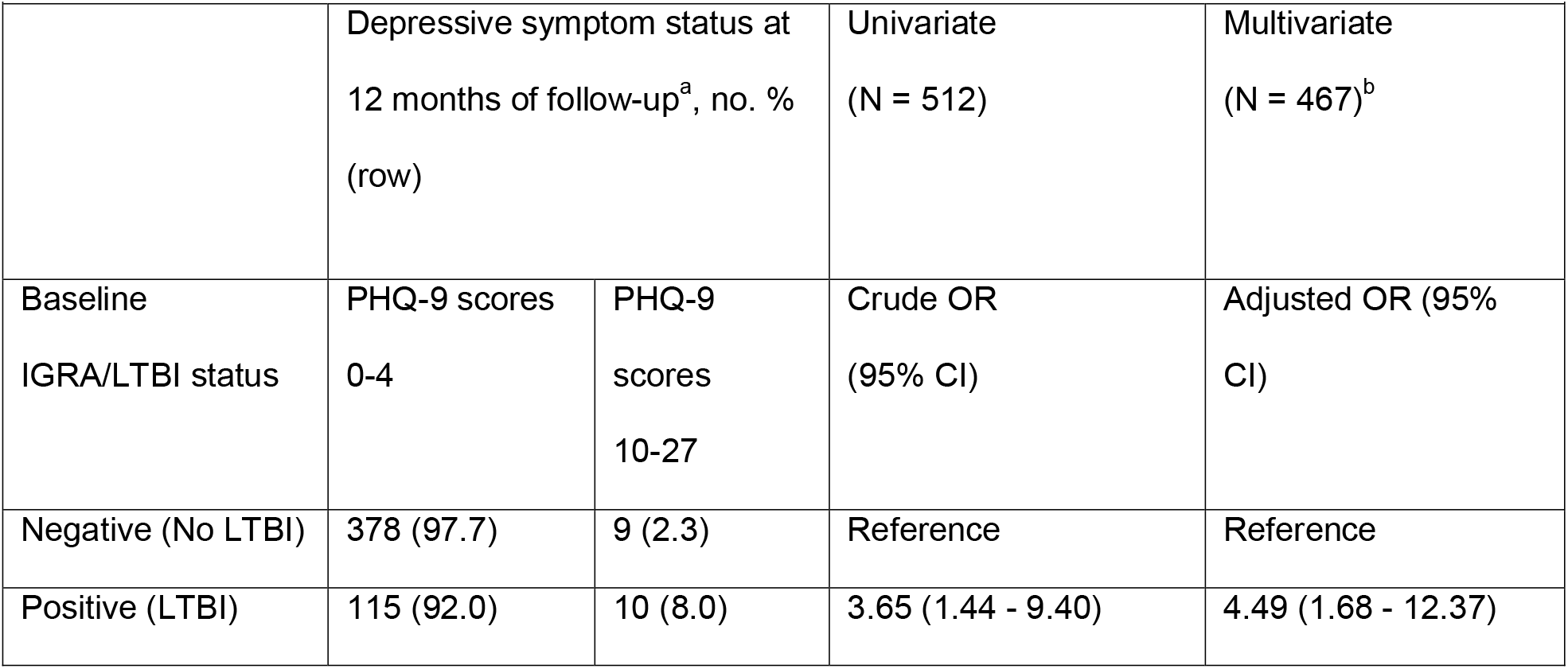
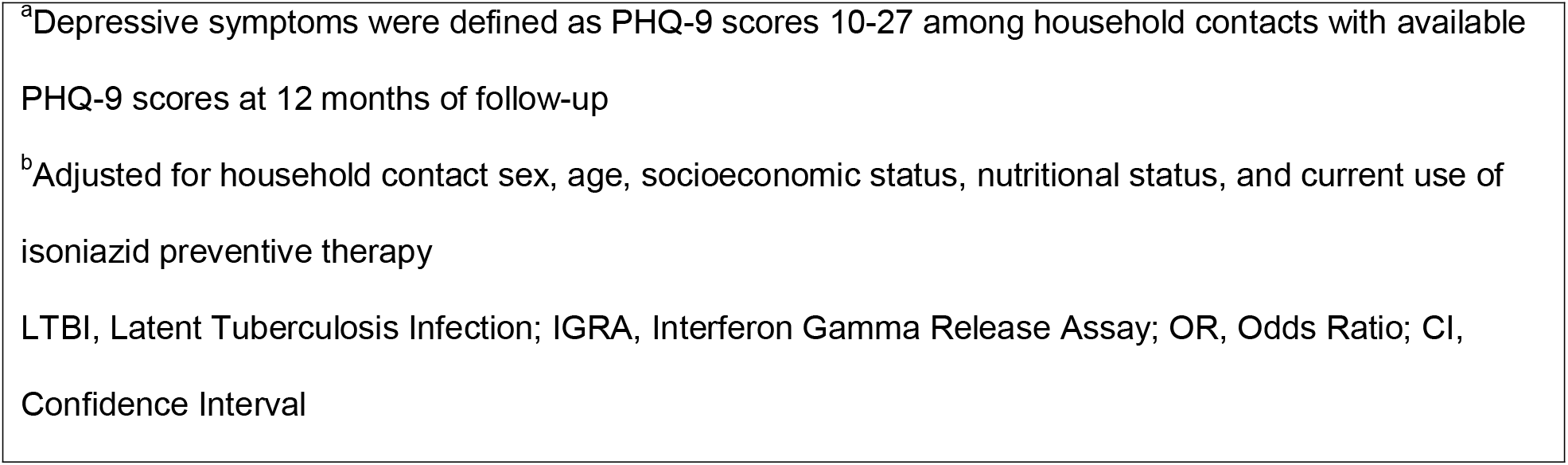
Association between baseline LTBI and depressive symptoms at 12 months of follow-up among HHCs with PHQ-9 scores 10-27 vs. 0-4.

|  | Depressive symptom status at 12 months of follow-up<sup>a</sup>, no. %<br>(row) |  | Univariate<br>(N = 512) | Multivariate<br>(N = 467)<sup>b</sup> |
| --- | --- | --- | --- | --- |
| Baseline IGRA/LTBI status | PHQ-9 scores 0-4 | PHQ-9 scores 10-27 | Crude OR (95% CI) | Adjusted OR (95% CI) |
| Negative (No LTBI) | 378 (97.7) | 9 (2.3) | Reference | Reference |
| Positive (LTBI) | 115 (92.0) | 10 (8.0) | 3.65 (1.44 - 9.40) | 4.49 (1.68 - 12.37) |

<sup>a</sup>Depressive symptoms were defined as PHQ-9 scores 10-27 among household contacts with available PHQ-9 scores at 12 months of follow-up
<sup>b</sup>Adjusted for household contact sex, age, socioeconomic status, nutritional status, and current use of isoniazid preventive therapy
LTBI, Latent Tuberculosis Infection; IGRA, Interferon Gamma Release Assay; OR, Odds Ratio; CI, Confidence Interval

## DISCUSSION

Here, we report that HHCs with LTBI at baseline had increased odds of experiencing depressive symptoms 12 months later. We also found that this association was driven by higher severities of depressive symptoms (PHQ-9 ≥10). We observed similar findings and trends when we further adjusted for incident TB disease status, restricted our analysis to those who recently acquired LTBI during follow-up, and used PHQ-9 ≥10 as an alternate cut-off.

Few studies have evaluated depression among close contacts of persons with TB specifically. Cross-sectional studies conducted in India and Nigeria have previously reported depression prevalence estimates ranging from 13.4% to 46.7% among contacts of people being treated for TB.^19,20^ In a large population-based cohort study in Taiwan, Pan et al. found no statistically significant difference in the incidence rate of depressive symptoms between HHCs of people being treated for TB and matched controls during a 5-year follow-up period. Further subgroup analysis revealed that female contacts had an increased risk of developing depressive symptoms within the first two years following time of index patient TB diagnosis.^12^ However, none of these observational studies evaluated the LTBI status of contacts. To our knowledge, no study has directly evaluated the association between LTBI and depressive symptoms among contacts of persons with TB.

We considered several possible explanations for the association between baseline LTBI and depressive symptoms at follow-up. First, depression is highly prevalent among persons with TB disease.^4^ HHCs represent an important group at elevated risk for developing active TB, which may precipitate mental conditions like depression. However, our data do not support this conclusion as the association between LTBI and depressive symptoms was not different among those who developed incident TB and those who did not. Second, we considered the possibility that there may be underlying biological mechanisms by which LTBI promotes the development of depressive symptoms.

Previous reports have indicated that host immune responses involved in LTBI may be considered a state of chronic, low-grade inflammation,^21–23^ which has been extensively studied and reviewed as a hypothesized pathophysiological mechanism for depression.^24–29^ For example, pro-inflammatory cytokines like interferon-gamma and tumor necrosis factor alpha (TNF-α) are known to be involved in activating and mediating the innate and adaptive immune response (e.g., monocyte/macrophage activation) to *M. tuberculosis* infection.^21–23^ The use of TNF-α inhibitors have been linked to an increased risk of reactivation of LTBI in clinical and animal studies, suggesting that TNF-α is a critical pro-inflammatory cytokine in immune-mediated processes like granuloma formation and maintenance.^22,23^ However, it remains unclear whether LTBI directly or indirectly contributes to the development of depression via inflammation-related mechanisms. Future human studies could consider measuring levels of inflammatory markers (e.g., IL-6 or CRP) and immune cells among individuals with LTBI and assess whether those markers change over time and are associated with a higher incidence of depression later. Further *in vitro*, animal, and clinical studies exploring the specific immunological roles that LTBI may play in the development of depression and other mental disorders are also needed. Third, our observed findings may be explained by various complex psychosocial factors such as stigmatization related to the diagnosis of LTBI itself, received stigma for being related or associated to a person with TB, fear, and psychological distress related to future uncertainties or transmitting TB to others.^30–34^

Our findings have several implications for the delivery of mental health care in TB programs. HHCs of persons being treated for TB represent an at-risk population for developing not only active TB but also mental conditions like depression and anxiety. Given that comorbid depression is associated with an increased risk for poor TB treatment outcomes,^6^ early intervention could attenuate progression to active TB and improve TB treatment outcomes for those who ultimately progress. Incorporating mental health screening for close contacts of individuals with TB, particularly those with LTBI, could be an effective way of detecting depressive symptoms early on and providing appropriate mental health services alongside routine preventive TB therapy. Ultimately, this could be one important form by which mental health care is integrated into TB programs.^35^

Our study had a couple limitations. First, as the PHQ-9 was limited to assessing depressive symptoms for the previous two weeks, we may have missed HHCs who were exhibiting depressive symptoms but were not captured by the PHQ-9 during follow-up. To address this, we included HHCs with available PHQ-9 data assessed at 12 months of follow-up with a two-month buffer period (10-14 months) to increase the total number of HHCs with potential depressive symptoms in our analytic sample. Second, there was a notable proportion of missing PHQ-9 data at follow-up (38.9%). This was partly due to initial hesitancy by study staff in asking questions regarding depressive symptoms to HHCs due to the perceived invasiveness of collecting sensitive mental health information; once detected, staff received re-training, but some data were lost. Nonetheless, we found no statistically significant difference in HHC baseline characteristics when comparing the complete/analytic and missing samples.

In conclusion, we found that HHCs with LTBI at baseline were more likely to develop depressive symptoms, and more severe symptoms, at 12 months of follow-up than those without LTBI at baseline. While further research is required to disentangle possible mechanisms behind this association, these findings suggest that close contacts of individuals with TB may represent an important population for both TB and mental health screening and care.

## Data Availability

All data produced in the present work are contained in the manuscript

## ACKNOWLEDGEMENTS

We acknowledge and thank the men and women who participated in this study, the Socios En Salud Sucursal Peru research personnel who conducted the home visits, and the health promoters working daily to assist people receiving TB treatment.

## AUTHOR’S CONTRIBUTIONS

JG, ALC, AS, CCH, and MM conceived of and designed the presented analysis. ALC performed the analysis. JG and ALC drafted the primary draft of the manuscript with input from all authors. JG, ALC, AS, JJ, RY, RG, ZZ, CCH, LL, and MM reviewed and revised subsequent drafts of the manuscript. All authors revised and approved the final version of the manuscript for submission.

## REFERENCES

1. Whiteford H, Ferrari A, Degenhardt L. Global Burden of Disease studies: Implications for mental and substance use disorders. Health Aff 2016;35:1114–20.

2. GBD 2019 Viewpoint Collaborators. Global burden of 369 diseases and injuries in 204 countries and territories, 1990–2019: a systematic analysis for the Global Burden of Disease Study 2019. The Lancet. 2020 Oct;396:1204–22.

3. World Health Organization. Global tuberculosis report 2022. Geneva, Switzerland: WHO, 2022. https://apps.who.int/iris/handle/10665/363752. Accessed October 2022.

4. Duko B, Bedaso A, Ayano G. The prevalence of depression among patients with tuberculosis: a systematic review and meta-analysis. Ann Gen Psychiatry. 2020;19:30.

5. Sweetland A, Oquendo M, Wickramaratne P, et al. Depression: a silent driver of the global tuberculosis epidemic. World Psychiatry. 2014 Oct;13:325–6.

6. Ruiz-Grosso P, Cachay R, de la Flor A, et al. Association between tuberculosis and depression on negative outcomes of tuberculosis treatment: A systematic review and meta-analysis. PloS One. 2020;15(1):e0227472.

7. Galea JT, Monedero-Recuero I, Sweetland AC. Beyond screening: a call for the routine integration of mental health care with tuberculosis treatment. Public Health Action. 2019 Mar 21;9(1):2.

8. World Health Organization. Guidance for national strategic planning for tuberculosis [Internet]. Geneva: World Health Organization; 2022. https://apps.who.int/iris/handle/10665/361418. Accessed November 2022.

9. Holmes KK, Bertozzi S, Bloom BR, et al. Disease Control Priorities, Third Edition (Volume 6): Major Infectious Diseases. The World Bank; 2017. http://elibrary.worldbank.org/doi/book/10.1596/978-1-4648-0524-0. Accessed November 2022.

10. Kiazyk S, Ball T. Latent tuberculosis infection: An overview. Can Commun Dis Rep. 2017 Mar 2;43:62–6.

11. Wong YJ, Noordin NM, Keshavjee S, et al. Impact of latent tuberculosis infection on health and wellbeing: a systematic review and meta-analysis. Eur Respir Rev. 2021 Mar 31;30(159):200260.

12. Pan SW, Yen YF, Feng JY, et al. The risk of depressive disorder among contacts of tuberculosis patients in a TB-endemic area: A population-based cohort study. Medicine (Baltimore). 2015 Oct;94(43):e1870.

13. Courtwright A, Turner AN. Tuberculosis and stigmatization: pathways and interventions. Public Health Rep Wash DC 1974. 2010 Aug;125 Suppl 4:34–42.

14. Khalaila R, Cohen M. Emotional suppression, caregiving burden, mastery, coping strategies and mental health in spousal caregivers. Aging Ment Health. 2016 Sep;20(9):908–17.

15. Oshio T. How is an informal caregiver’s psychological distress associated with prolonged caregiving? Evidence from a six-wave panel survey in Japan. Qual Life Res. 2015 Dec;24(12):2907–15.

16. Moody DB, Murray MB. Metabolic factors that control the spectrum of human tuberculosis. Granttome; 2015. https://grantome.com/grant/NIH/U19-AI111224-07. Accessed November 2022.

17. Calderón M, Gálvez-Buccollini JA, Cueva G, et al. Validación de la versión peruana del PHQ-9 para el diagnóstico de depresión. Rev Peru Med Exp Salud Pública. 2012 Dec;29(4):578–9.

18. Kroenke K, Spitzer RL, Williams JB. The PHQ-9: validity of a brief depression severity measure. J Gen Intern Med. 2001 Sep;16(9):606–13.

19. Ige OM, Lasebikan VO. Prevalence of depression in tuberculosis patients in comparison with non-tuberculosis family contacts visiting the DOTS clinic in a Nigerian tertiary care hospital and its correlation with disease pattern. Ment Health Fam Med. 2011 Dec;8(4):235–41.

20. Itagi ABH, Dharmalingam A, Yunus GY, et al. Comparative assessment of depression, quality of sleep, and respiratory functions among tuberculosis patients with their nontuberculosis family contacts. Indian J Respir Care. 2021;10(3):6.

21. Mack U, Migliori GB, Sester M, et al. LTBI: latent tuberculosis infection or lasting immune responses to M. tuberculosis? A TBNET consensus statement. Eur Respir J. 2009 May;33(5):956–73.

22. Lin PL, Flynn JL. Understanding latent tuberculosis: a moving target. J Immunol Baltim Md 1950. 2010 Jul 1;185(1):15–22.

23. Dutta NK, Karakousis PC. Latent tuberculosis infection: myths, models, and molecular mechanisms. Microbiol Mol Biol Rev MMBR. 2014 Sep;78(3):343–71.

24. Dantzer R, O’Connor JC, Freund GG, et al. From inflammation to sickness and depression: when the immune system subjugates the brain. Nat Rev Neurosci. 2008 Jan;9(1):46–56.

25. Maes M. Depression is an inflammatory disease, but cell-mediated immune activation is the key component of depression. Prog Neuropsychopharmacol Biol Psychiatry. 2011 Apr 29;35(3):664–75.

26. Khandaker GM, Dantzer R, Jones PB. Immunopsychiatry: important facts. Psychol Med. 2017 Oct;47(13):2229–37.

27. Leonard BE. Inflammation and depression: a causal or coincidental link to the pathophysiology? Acta Neuropsychiatr. 2018 Feb;30(1):1–16.

28. Bullmore E. The art of medicine: Inflamed depression. Lancet Lond Engl. 2018 Oct 6;392(10154):1189–90.

29. Beurel E, Toups M, Nemeroff CB. The bidirectional relationship of depression and inflammation: Double trouble. Neuron. 2020 Jul 22;107(2):234–56.

30. Coreil J, Lauzardo M, Clayton H. Stigma and therapy completion for latent tuberculosis among Haitian-origin patients. Fla Public Health Rev. 2010 Jan;7:32–8.

31. Brown J, Capocci S, Smith C, et al. Health status and quality of life in tuberculosis. Int J Infect Dis. 2015 Mar;32:68–75.

32. Mason PH, Degeling C, Denholm J. Sociocultural dimensions of tuberculosis: an overview of key concepts. Int J Tuberc Lung Dis. 2015 Oct 1;19(10):1135–43.

33. Fox GJ, Dobler CC, Marais BJ, et al. Preventive therapy for latent tuberculosis infection—the promise and the challenges. Int J Infect Dis. 2017 Mar;56:68–76.

34. Shedrawy J, Jansson L, Röhl I, et al. Quality of life of patients on treatment for latent tuberculosis infection: a mixed-method study in Stockholm, Sweden. Health Qual Life Outcomes. 2019 Oct 24;17(1):158.

35. Sweetland AC, Jaramillo E, Wainberg ML, et al. Tuberculosis: an opportunity to integrate mental health services in primary care in low-resource settings. Lancet Psychiatry. 2018 Dec;5(12):952–4.

