## Supplemental Tables for "Latent tuberculosis and depressive symptoms in household contacts of persons with active TB: A cohort study"

| **Supplementary Table 1. Comparison of the complete/analytic and missing data samples** | | | |
| --- | --- | --- | --- |
|  | Analytic sample vs. missing data sample, no. % (column) | |  |
| Household contact characteristic | Complete/analytic sample^a^ (n = 563) | Missing data sample^b^ (n = 358) | p-value^c^ |
| Age (years) |  |  |  |
| 12 - 24 | 182 (32.3) | 100 (27.9) | 0.364 |
| 24 - 48 | 242 (43.0) | 162 (45.3) |  |
| >48 | 139 (24.7) | 96 (26.8) |  |
| Male sex | 237 (42.1) | 146 (40.8) | 0.745 |
| Smoking status |  |  |  |
| Non-smoker | 522 (92.7) | 335 (93.6) | 0.714 |
| Smoker | 41 (7.3) | 23 (6.4) |  |
| Alcohol intake status |  |  |  |
| Non-drinker | 422 (75.0) | 272 (76.0) | 0.785 |
| Drinker | 141 (25.0) | 86 (24.0) |  |
| HIV seropositive | 3 (0.5) | 2 (0.6) | 1.000 |
| Heart Disease | 11 (2.0) | 4 (1.1) | 0.427 |
| Diabetic | 22 (4.0) | 14 (3.9) | 1.000 |
| Socioeconomic status |  |  |  |
| Low | 173 (30.7) | 106 (29.7) | 0.614 |
| Medium | 217 (38.5) | 149 (41.7) |  |
| High | 173 (30.7) | 102 (28.6) |  |
| Nutritional status |  |  | 0.066 |
| Normal | 182 (35.3) | 48 (26.7) |  |
| Overweight | 332 (64.3) | 131 (72.8) |  |
| Underweight | 2 (0.4) | 1 (0.5) |  |
| Current use of IPT | 8 (1.4) | 3 (0.8) | 0.543 |
| ^a^ Sample of household contacts with both baseline IGRA results and depressive symptom data at 12 months of follow-up  ^b^ Sample of household contacts with only baseline IGRA results (i.e., missing depressive symptom data at 12 months of follow-up)  ^c^ p-values were obtained using the Chi-square test or Fisher’s Exact tests for cells with $\leq$5 observations  IGRA, Interferon Gamma Release Assay; HIV, Human Immunodeficiency Virus; IPT, Isoniazid Preventive Therapy | | | |

| **Supplementary Table 2. Association between baseline LTBI and depressive symptoms at 12 months of follow-up among household contacts with IGRA results at baseline and 12 months of follow-up** | | | | |
| --- | --- | --- | --- | --- |
|  | Depressive symptom status at 12 months of follow-up^a^, no. (row %) | | Univariate  (N = 418) | Multivariate  (N = 388)^b^ |
| IGRA Conversion Status | PHQ-9 scores 0-4 | PHQ-9 scores 5-27 | Crude OR (95% CI) | Adjusted OR (95% CI) |
| No IGRA conversion (i.e., negative IGRA result at baseline and negative IGRA result at follow-up) | 329 (90.1) | 36 (9.9) | Reference | Reference |
| IGRA conversion (i.e., negative IGRA result at baseline and positive IGRA results at follow-up) | 45 (84.9) | 8 (15.1) | 1.62 (0.67 - 3.56) | 1.60 (0.63 - 3.67) |
| ^a^ Depressive symptoms were defined as PHQ-9 scores 5-27 among household contacts with available PHQ-9 scores at 12 months of follow-up  ^b^ Adjusted for household contact sex, age, socioeconomic status, nutritional status, and current use of isoniazid preventive therapy  LTBI, Latent Tuberculosis Infection; IGRA, Interferon Gamma Release Assay; OR, Odds Ratio; CI, Confidence Interval | | | | |

| **Supplementary Table 3. Association between baseline LTBI and depressive symptoms at 12 months of follow-up among household contacts using PHQ-9** $\boldsymbol{\geq}$**10 to define depressive symptoms** | | | | |
| --- | --- | --- | --- | --- |
|  | Depressive symptom status at 12 months of follow-up^a^, no. (Row %) | | Univariate  (N = 563) | Multivariate  (N = 516)^b^ |
| Baseline IGRA/LTBI status | PHQ-9 scores 0-9 | PHQ-9 scores 10-27 | Crude OR (95% CI) | Adjusted OR (95% CI) |
| Negative (No LTBI) | 413 (97.9) | 9 (2.1) | Reference | Reference |
| Positive (LTBI) | 131 (92.9) | 10 (7.1) | 3.50 (1.38 - 9.00) | 4.30 (1.61 - 11.90) |
| ^a^ Depressive symptoms were defined as PHQ-9 scores 10-27 among household contacts with available PHQ-9 scores at 12 months of follow-up  ^b^ Adjusted for household contact sex, age, socioeconomic status, nutritional status, and current use of isoniazid preventive therapy  LTBI, Latent Tuberculosis Infection; IGRA, Interferon Gamma Release Assay; OR, Odds Ratio; CI, Confidence Interval | | | | |
